# Malnutrition and Micronutrient Deficiency among Pregnant Women under Pradhan Mantri Surakshit Matritva Abhiyan in Jharkhand, India: A Cross-sectional Study

**DOI:** 10.64898/2026.08.19.26360853

**Authors:** Archana Kumari, Kumari Asha Kiran, Shailesh Sanjul Hembrom, Manisha Kujur, Ratnesh Sinha, Anuj Kumar Anit

## Abstract

**Objective:** Malnutrition and micronutrient deficiency are significant public health problems affecting the well-being of both the mother and her offspring. It is important to first quantify their burden in underserved communities and then to tackle this problem. The objective of this study was to assess the nutritional status, micronutrient deficiency profile, and associated determinants among pregnant women attending antenatal care clinics under PMSMA in selected government health facilities across three districts of Jharkhand, India.

**Design:** The study employed a cross-sectional study design to assess the burden of malnutrition and micronutrient deficiency among pregnant women.

**Setting:** The study was conducted in 4 health facilities in 3 districts of Jharkhand. The selected facilities were Rajendra Institute of Medical Sciences (RIMS), Ranchi; District Hospital (Sadar), Ranchi; District Hospital (Sadar), Godda; and Community Health Centre (Gamharia, Saraikela) under RHTC, Department of Community Medicine, Manipal Tata Medical College, Jamshedpur. The study was conducted during the period of September to December, 2023.

**Participant:** Eligible pregnant women attending ANC clinics under PMSMA in the selected health facilities were enrolled until the required sample size of 977 was achieved. Pregnant women who were critically ill or those who presented with some emergency conditions were excluded from the study.

**Result:** Based on BMI assessment, 38.6% of participants were malnourished, with 17.7% being underweight and 20.9% overweight or obese. Anaemia affected 69.3% of women, while clinical features suggestive of iron deficiency were observed among 21.1% of the participants. Vitamin A deficiency, iodine deficiency, and fluoride excess were also identified among a smaller proportion of women.

**Conclusion:** The present study highlights a substantial burden of both malnutrition and micronutrient deficiency among pregnant women in Jharkhand and justifies the need for integrated maternal nutrition strategies during antenatal care.

## Introduction

Maternal nutrition is a critical determinant of pregnancy outcomes and early growth and development of the offspring. Malnutrition, defined as an imbalance in energy and/or nutrient intake, includes both undernutrition and overnutrition and assumes particular importance during pregnancy because maternal nutrition supports both maternal physiological needs and foetal growth and development. [1,2] A state of undernutrition during pregnancy can compromise maternal health, increase obstetric risk, and contribute to adverse neonatal outcomes such as low birth weight, small for gestational age and preterm birth. On the other hand, overnutrition during pregnancy is associated with complications such as gestational diabetes, hypertensive disorders, postpartum haemorrhage and congenital anomalies. [3] Therefore, assessment of nutritional status during pregnancy should be an essential component of antenatal care.

Globally, populations are increasingly experiencing the double burden of malnutrition, characterized by the coexistence of undernutrition and overnutrition within the same population. [4] This transition is driven by changing dietary patterns, increased consumption of packaged and processed foods, lifestyle changes, urbanization, and increased exposure to conditions which have been termed as obesogenic environments. [5,6] Body Mass Index, although widely used for nutritional assessment, has limitations during pregnancy because gestational weight gain alters interpretation. While some Indian antenatal care guidelines permit BMI use before 20 weeks of gestation, other authorities such as the Institute of Medicine (IOM), USA emphasize trimester-wise weight gain based on pre-pregnancy BMI. [7–9] Nevertheless, BMI remains a practical screening tool in resource-limited settings where repeated nutritional surveillance may be difficult.

Alongside, anthropometric malnutrition and micronutrient deficiencies remain a major concern during pregnancy. Adequate micronutrient status is essential for normal physiological function, maternal well-being, placental function, and foetal development. Deficiencies of iron, folate, vitamin A, iodine, and other micronutrients are common during pregnancy and are often described as “hidden hunger” because they may occur without obvious clinical manifestations but can substantially affect maternal and child health. [10–12] Anaemia, commonly due to iron deficiency, affects an estimated 40% of pregnant women globally, with the highest burden in South East Asia, Africa, and the Eastern Mediterranean regions. [13] Vitamin A deficiency, iodine deficiency, and concerns related to fluoride exposure during pregnancy further highlight the need to assess micronutrient status in pregnant women. [14–18]

In India, maternal mortality remains a continuing public health priority. The National Family Health Survey data shows that 43% of women of reproductive age have either undernutrition or overnutrition based on their BMI, while the corresponding figure for Jharkhand is 38.1%. [19] NFHS-5 data for Jharkhand also reflects suboptimal antenatal care and supplementation coverage, with only 39% of pregnant women completing 4 antenatal visits, 28% consuming iron-folic acid tablets for 100 days or more, and 15% consuming them for 180 days or more. The burden is further underscored by the high prevalence of anemia among pregnant women in the state. These indicators are especially relevant in Jharkhand, where rural-urban disparities, social vulnerability, and dietary inadequacy may contribute simultaneously to undernutrition, overnutrition, and micronutrient deficiency.

Several national and state-level initiatives, including the Integrated Child Development Services and Pradhan Mantri Surakshit Matritva Abhiyan (PMSMA), aim to improve maternal nutrition and strengthen antenatal care services. PMSMA provides a strategic platform for screening, early detection, referral, and management of high-risk pregnancies, thereby contributing to reduction of maternal and neonatal morbidity and mortality. [20] However, evidence on the combined burden of anthropometric malnutrition and micronutrient deficiency among pregnant women attending public antenatal clinics in Jharkhand remains limited.

The present study was undertaken to assess the nutritional status, micronutrient deficiency profile, and associated determinants among pregnant women attending antenatal care clinics under Pradhan Mantri Surakshit Matritva Abhiyan (PMSMA) in selected government health facilities across three districts of Jharkhand.

## Methods

This was a cross-sectional study conducted in 4 health facilities in 3 districts of Jharkhand. The selected facilities were Rajendra Institute of Medical Sciences (RIMS), Ranchi; District Hospital (Sadar), Ranchi; District Hospital (Sadar), Godda; and Community Health Centre (Gamharia, Saraikela) under RHTC, Department of Community Medicine, Manipal Tata Medical College, Jamshedpur. The study was conducted during the period of September to December, 2023.

A consecutive sampling method was employed and eligible pregnant women attending ANC clinics under PMSMA in the selected health facilities during the study period were enrolled until the required sample size was achieved. Pregnant women who were critically ill or those who presented with some emergency condition during the period of data collection were excluded from the study. Informed consent was taken before enrolling participants for the study.

### Sample size

Sample size for the study was calculated by the given formula:

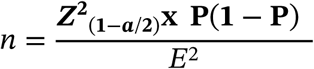

| **Prevalence of an indicator (P)** | **District Hospital, Ranchi** | **RIMS, Ranchi** | **District Hospital, Godda** | **CHC Gamharia, Saraikela Kharsawan** |  |
|---|---|---|---|---|---|
| Women whose Body Mass Index (BMI) is below normal (BMI <18.5 kg/m<sup>2</sup>) (%) | 17.3% (urban) | 17.3% (urban) | 17.3% (urban) | 29.2% (rural) | Total Sample 977.1 (Approximately 977) |
| Level of Significance (α) | 5% | 5% | 5% | 5% |  |
| Z-score (Zα) | 1.96 | 1.96 | 1.96 | 1.96 |  |
| Desired precision (E) | 5% | 5% | 5% | 5% |  |
| Required minimum sample size (n) | 219.8 | 219.8 | 219.8 | 317.7 |  |

Based on recent NFHS 5 (Jharkhand) estimates, women whose BMI is below normal <18.5 kg/m^2^ (according to area wise calculation), the total sample size was taken as 977.

### Data Collection Procedure

After taking consent, data was collected by face-to-face interview using a structured and pretested questionnaire. The interview was taken after the pregnant women had received the ANC services. Each pregnant woman was interviewed privately and confidentiality was maintained. Height, weight and body mass index (BMI) of the study participants was calculated using pre-tested and calibrated instruments. Data collectors (counsellor/ staff nurse/ ANM/ residents) and pregnant women were informed to follow the infection prevention protocols (if required). The filled questionnaires were collected and checked for consistency and missing data by investigators. Any missing data which could be collected by telephonic conversation was done by contacting the participants by telephone. Responses missing information to be collected physically such as height and weight were discarded.

Clinical assessment of the participants was done to collect data on anaemia, vitamin A, and iodine deficiency and fluoride excess. anaemia was assessed on the basis of haemoglobin levels of the participants. Vitamin A deficiency was assessed on the basis of presence or absence of clinical symptoms and signs, namely night blindness or Bitot’s spots, respectively. Iodine deficiency was evaluated on the basis of presence or absence of goitre. Fluoride excess was assessed on the basis of presence or absence of clinical signs and symptoms suggestive of dental or skeletal fluorosis. Clinical anaemia was determined on the basis of clinical features such as dizziness, weakness and breathlessness and presence of pallor.

Dietary assessment was done using a food frequency questionnaire with the food groups categorized as per Minimum Dietary Diversity for Women (MDD-W) indicator. [21]

### Data Analysis

The baseline survey data was collected in Google forms and transferred to MS Office Excel spreadsheet. Descriptive data analysis was done using Jamovi software version 2.3. Continuous data was presented as mean with standard deviation. Categorical data was described in terms of proportion. Chi-square test was done to check the association between the independent and dependent variables. Multinomial logistic regression was done to calculate the adjusted odds ratio and estimate the risk. A ‘p value’ less than 0.05 was considered as statistically significant.

### Ethics approval

Ethical clearance was obtained from Institutional Ethics Committee, Rajendra Institute of Medical Sciences, Ranchi (Letter no. 65, dated 09.03.2023) prior to the conduct of the study.

Patient and public involvement: Patients or the public were not involved in the design, or conduct, or reporting, or dissemination plans of our research.

## Results

### Socio-demographic characteristics

A total of 977 pregnant women participated in the study. Most of the participants were aged 20- 24 years (43.8%), followed by 25-29 years (30.5%), while 13.7% were below 20 years. Majority of the participants belonged to families with fewer than five members (64.3%). Hindus constituted 70.5% of the study population, followed by Muslims (17.9%). In terms of category distribution, 43.3% belonged to other backward classes, 21.3% to scheduled tribes, and 19.8% to the general category. Regarding education, 12.3% were illiterate, while the remaining had varying levels of formal education, with 22.7% educated up to the intermediate level and 20.7% graduates. The vast majority were housewives (94.1%). Among husbands, 57.0% were daily wage labourers. Nearly equal distribution of participants was observed across income groups, with 49.5% having a monthly family income below Rs. 10,000. District-wise, 57.9% of participants were recruited from Ranchi, 27.6% from Godda and 14.4% from Saraikela. (Table 1)

**Table 1:** Socio-demographic characteristics, obstetric profile, laboratory and clinical findings and dietary assessment of the study participants (n=977)

| Socio-demographic variables |  | Frequency | Percentage |
|---|---|---|---|
| Age group | <20 years | 134 | 13.7 |
|  | 20-24 years | 428 | 43.8 |
|  | 25-29 years | 298 | 30.5 |
|  | >29 years | 117 | 12.0 |
| Number of family members | <5 | 628 | 64.3 |
|  | ≥5 | 349 | 35.7 |
| Religion | Hindu | 689 | 70.5 |
|  | Muslim | 175 | 17.9 |
|  | Christian | 35 | 3.6 |
|  | Others | 78 | 8.0 |
| Category | General | 193 | 19.8 |
|  | Scheduled Caste | 76 | 7.8 |
|  | Scheduled Tribe | 208 | 21.3 |
|  | Other Backward Classes | 423 | 43.3 |
|  | Others | 77 | 7.9 |
| Educational status | Illiterate | 120 | 12.3 |
|  | Primary | 218 | 22.3 |
|  | Middle | 215 | 22.0 |
|  | Inter | 222 | 22.7 |
|  | Graduate | 202 | 20.7 |
| Occupation | Salaried employee | 21 | 2.1 |
|  | Housewife | 919 | 94.1 |
|  | Daily wage labour | 22 | 2.3 |
|  | Household helper | 4 | 0.4 |
|  | Business | 11 | 1.1 |
| Husband's Occupation | Salaried employee | 178 | 18.2 |
|  | Daily wage labour | 557 | 57.0 |
|  | Unemployed | 33 | 3.4 |
|  | Business | 209 | 21.4 |
| District of enrolment | Ranchi | 566 | 57.9 |
|  | Godda | 270 | 27.6 |
|  | Saraikela | 141 | 14.4 |
| Monthly family income (in ₹) | <10000 | 484 | 49.5 |
|  | ≥10000 | 493 | 50.5 |
| **Obstetric profile of study participants** |  |  |  |
| Number of ANC checkups | <3 | 369 | 37.8 |
|  | ≥3 | 608 | 62.2 |
| Presenting trimester | First trimester | 73 | 7.5 |
|  | Second trimester | 408 | 41.8 |
|  | Third trimester | 496 | 50.8 |
| Gravida | Primigravida | 492 | 50.4 |
|  | Multigravida | 485 | 49.6 |
| History of abortions | No | 787 | 80.6 |
|  | Yes | 190 | 19.4 |
| **Laboratory and clinical findings** |  |  |  |
| Hb levels | Normal (≥11 gm/dl) | 301 | 30.8 |
|  | Mild anaemia (10-10.9 gm/dl) | 370 | 37.9 |
|  | Moderate anaemia (7-9.9 gm/dl) | 278 | 28.4 |
|  | Severe anaemia (<7 gm/dl) | 28 | 2.9 |
| Vitamin A deficiency | No | 965 | 98.8 |
|  | Yes | 12 | 1.2 |
| Iodine deficiency | No | 972 | 99.5 |
|  | Yes | 5 | 0.5 |
| Fluoride excess | No | 963 | 98.6 |
|  | Yes | 14 | 1.4 |
| Clinical anaemia | No | 771 | 78.9 |
|  | Yes | 206 | 21.1 |
| **Dietary assessment** |  |  |  |
| Frequency of meals per day | <3 | 217 | 22.2 |
|  | ≥3 | 760 | 77.8 |
| Frequency of grains, white roots and tubers, and plantains | Less than twice a week | 24 | 2.5 |
|  | Twice a week or more | 953 | 97.5 |
| Frequency of pulses | Less than twice a week | 103 | 10.5 |
|  | Twice a week or more | 874 | 89.5 |
| Frequency of nuts and seeds | Less than twice a week | 616 | 63.1 |
|  | Twice a week or more | 361 | 36.9 |
| Frequency of milk and milk products | Less than twice a week | 326 | 33.4 |
|  | Twice a week or more | 651 | 66.6 |
| Frequency of meat, poultry and fish | Less than twice a week | 862 | 88.2 |
|  | Twice a week or more | 115 | 11.8 |
| Frequency of egg | Less than twice a week | 864 | 88.4 |
|  | Twice a week or more | 113 | 11.6 |
| Frequency of dark green leafy vegetables | Less than twice a week | 285 | 29.2 |
|  | Twice a week or more | 692 | 70.8 |
| Frequency of other vitamin-A rich fruits and vegetables | Less than twice a week | 696 | 71.2 |
|  | Twice a week or more | 281 | 28.8 |
| Frequency of other vegetables | Less than twice a week | 84 | 8.6 |
|  | Twice a week or more | 893 | 91.4 |
| Frequency of other fruits | Less than twice a week | 295 | 30.2 |
|  | Twice a week or more | 682 | 69.8 |
| Any particular food item avoided during pregnancy? | Papaya | 572 | 58.5 |
|  | Meat, eggs and fish | 22 | 2.3 |
|  | Pineapple | 2 | 0.2 |

### Obstetric profile

Majority of the participants had completed at least three antenatal care visits (62.2%). Nearly half (50.8%) were in the third trimester at the time of assessment, while only 7.5% were in the first trimester. Participants were almost equally distributed between primigravida (50.4%) and multigravida (49.6%). A history of abortion was reported by 19.4% of participants. (Table 1)

### Laboratory and clinical findings

69.2% (676 out of 977) women were diagnosed with anaemia, with the proportions of mild, moderate and severe anaemia being 37.9%, 28.4% and 2.9% respectively of the total. (Table 1) 12 participants (1.2%) out of 977 presented with signs or symptoms of Vitamin A Deficiency. A total of 5 participants (0.5%) out of 977 presented with goitre. 14 of the women (1.3%) were diagnosed with fluorosis, 12 with dental fluorosis (1.1%) and 2 with skeletal fluorosis (0.2%). A total of 21.1% participants presented with clinical features suggestive of anaemia such as pallor, weakness and breathlessness, and dizziness. (Table 1)

### Dietary Assessment

Majority of women (77.8%) reported consuming at least 3 meals in a day. Grains, white roots and tubers, and plantains were the staple food of the majority of women with 97.5% women reporting twice weekly intakes. The consumption of pulses was lower with 89.5% women reporting at least twice weekly intake. Nuts and seeds, rich sources of proteins, fats and micronutrients held a meagre importance in the diets of the participants with around a third (36.9%) reporting at least twice weekly intake while 63.1% reporting an intake of less than two times in a week. Milk and milk products were part of frequent intake of nearly two-thirds (66.6%) households reporting at least twice weekly consumption. Meat, poultry and fish were reported to be consumed mostly on a weekly basis with 88.2% women reporting intakes of less than twice a week, which was similar to the consumption of eggs, with 88.4% of participants reporting the same. Dark green leafy vegetables, which are particularly rich sources of iron were subject to regular consumption, with 70.8% women reporting intake of twice or more in a week. A large proportion reported no or just weekly intake of vitamin-A rich fruits and vegetables (71.2%) such as papaya, carrot and pumpkin, reflecting a cause for concern regarding vitamin-A deficiency. Other vegetables were mostly consumed on a frequent basis with 91.4% households reporting intakes of twice or more in a week. The frequency of consuming other fruits was varied with nearly one-third of the participants (30.2%) reporting intakes of less than twice in a week while more than two-thirds (69.8%) reporting twice weekly consumption or more. (Table 1)

Some food items were specifically avoided during the course of pregnancy, such as papaya (with 58.5% of participants avoiding it), followed by meat, eggs, and fish (2.3%), and pineapple (0.2%). (Table 1)

### Prevalence of malnutrition

Based on WHO classification of BMI, 17.7% of participants were undernourished, 61.4% had normal nutritional status, while 20.9% were overnourished (overweight 17.4%, obese 3.5%). (Figure 1)

**Figure 1:**
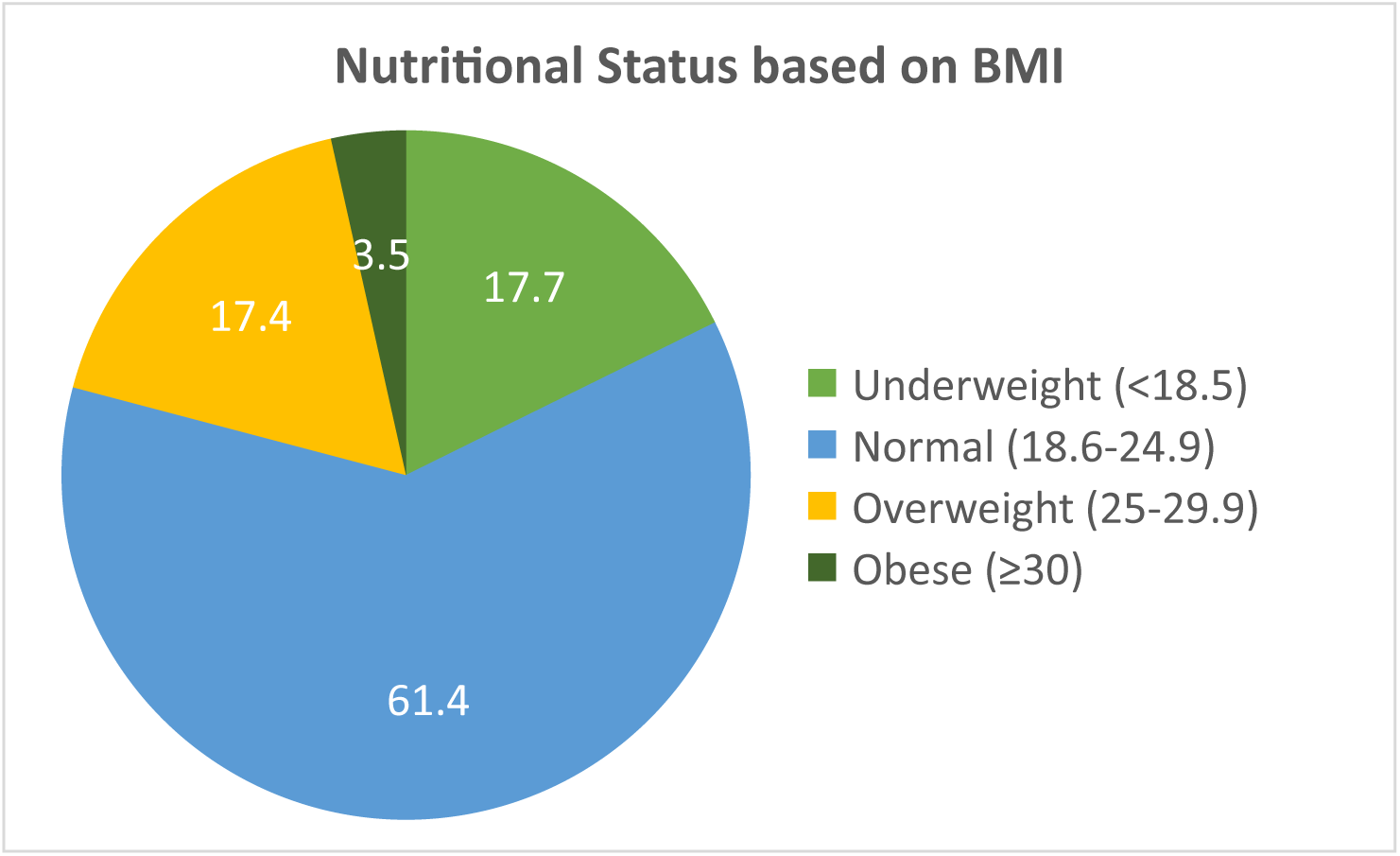
Pie chart showing distribution of participants according to their BMI (n=977)

### Association of sociodemographic and obstetric factors and laboratory and clinical findings with nutritional status

Significant associations were observed between BMI-based nutritional status and several variables.

Age groups showed a statistically significant association (p<0.001), with higher over-nutrition observed in older age groups. Category (p<0.001), educational status (p<0.001), occupation (p=0.001), and husband’s occupation (p<0.001) were also significantly associated with nutritional status. District-wise variation was significant (p<0.001), with higher undernutrition in Saraikela (31.9%) and higher overnutrition in Ranchi (28.3%). Monthly family income was also significantly associated (p<0.001), with higher over-nutrition among those with incomes greater than ₹10,000. (Table 2)

**Table 2:**
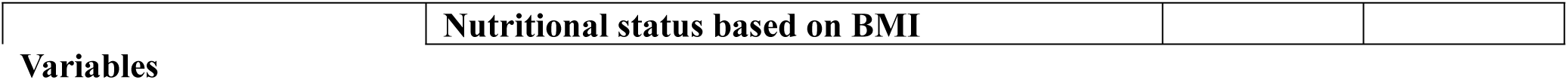
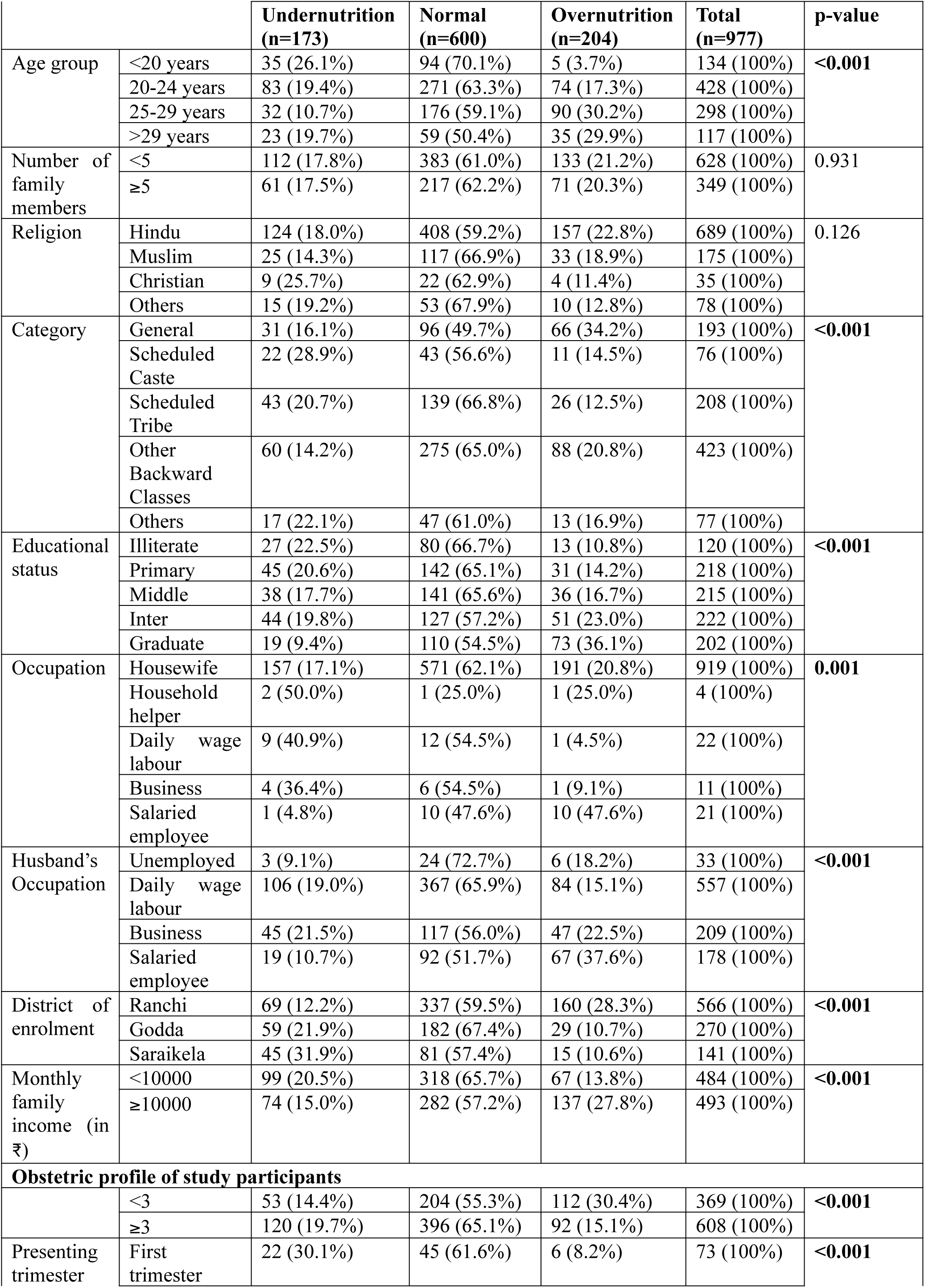
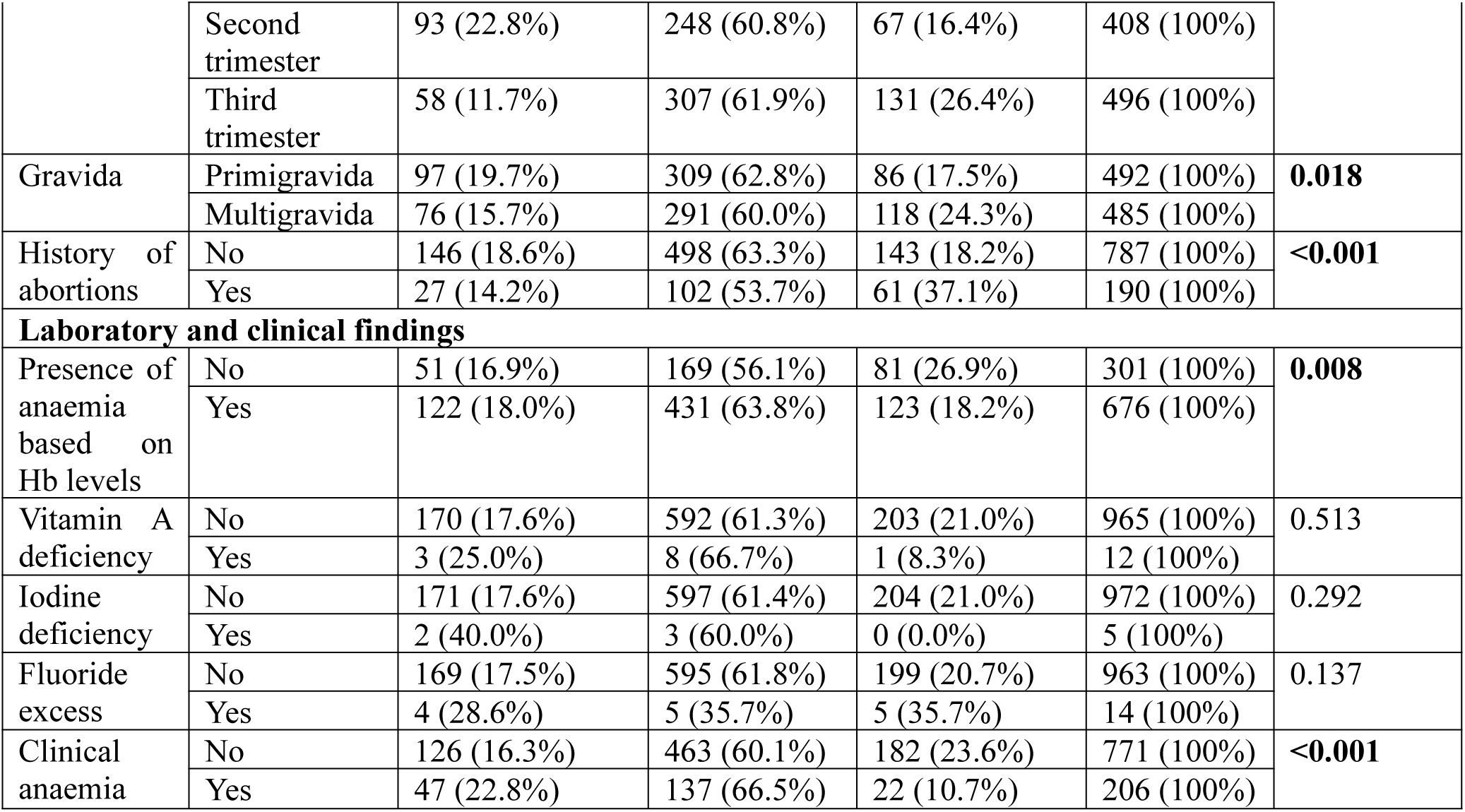
Chi-square test for association of socio-demographic and obstetric factors and laboratory and clinical findings with nutritional status

| **Variables** |  | **Nutritional status based on BMI** |  |  |  |  |
|---|---|---|---|---|---|---|
|  |  | **Undernutrition<br>(n=173)** | **Normal<br>(n=600)** | **Overnutrition<br>(n=204)** | **Total<br>(n=977)** | **p-value** |
| Age group | <20 years | 35 (26.1%) | 94 (70.1%) | 5 (3.7%) | 134 (100%) | **<0.001** |
|  | 20-24 years | 83 (19.4%) | 271 (63.3%) | 74 (17.3%) | 428 (100%) |  |
|  | 25-29 years | 32 (10.7%) | 176 (59.1%) | 90 (30.2%) | 298 (100%) |  |
|  | >29 years | 23 (19.7%) | 59 (50.4%) | 35 (29.9%) | 117 (100%) |  |
| Number of family members | <5 | 112 (17.8%) | 383 (61.0%) | 133 (21.2%) | 628 (100%) | 0.931 |
|  | ≥5 | 61 (17.5%) | 217 (62.2%) | 71 (20.3%) | 349 (100%) |  |
| Religion | Hindu | 124 (18.0%) | 408 (59.2%) | 157 (22.8%) | 689 (100%) | 0.126 |
|  | Muslim | 25 (14.3%) | 117 (66.9%) | 33 (18.9%) | 175 (100%) |  |
|  | Christian | 9 (25.7%) | 22 (62.9%) | 4 (11.4%) | 35 (100%) |  |
|  | Others | 15 (19.2%) | 53 (67.9%) | 10 (12.8%) | 78 (100%) |  |
| Category | General | 31 (16.1%) | 96 (49.7%) | 66 (34.2%) | 193 (100%) | **<0.001** |
|  | Scheduled Caste | 22 (28.9%) | 43 (56.6%) | 11 (14.5%) | 76 (100%) |  |
|  | Scheduled Tribe | 43 (20.7%) | 139 (66.8%) | 26 (12.5%) | 208 (100%) |  |
|  | Other Backward Classes | 60 (14.2%) | 275 (65.0%) | 88 (20.8%) | 423 (100%) |  |
|  | Others | 17 (22.1%) | 47 (61.0%) | 13 (16.9%) | 77 (100%) |  |
| Educational status | Illiterate | 27 (22.5%) | 80 (66.7%) | 13 (10.8%) | 120 (100%) | **<0.001** |
|  | Primary | 45 (20.6%) | 142 (65.1%) | 31 (14.2%) | 218 (100%) |  |
|  | Middle | 38 (17.7%) | 141 (65.6%) | 36 (16.7%) | 215 (100%) |  |
|  | Inter | 44 (19.8%) | 127 (57.2%) | 51 (23.0%) | 222 (100%) |  |
|  | Graduate | 19 (9.4%) | 110 (54.5%) | 73 (36.1%) | 202 (100%) |  |
| Occupation | Housewife | 157 (17.1%) | 571 (62.1%) | 191 (20.8%) | 919 (100%) | **0.001** |
|  | Household helper | 2 (50.0%) | 1 (25.0%) | 1 (25.0%) | 4 (100%) |  |
|  | Daily wage labour | 9 (40.9%) | 12 (54.5%) | 1 (4.5%) | 22 (100%) |  |
|  | Business | 4 (36.4%) | 6 (54.5%) | 1 (9.1%) | 11 (100%) |  |
|  | Salaried employee | 1 (4.8%) | 10 (47.6%) | 10 (47.6%) | 21 (100%) |  |
| Husband’s Occupation | Unemployed | 3 (9.1%) | 24 (72.7%) | 6 (18.2%) | 33 (100%) | **<0.001** |
|  | Daily wage labour | 106 (19.0%) | 367 (65.9%) | 84 (15.1%) | 557 (100%) |  |
|  | Business | 45 (21.5%) | 117 (56.0%) | 47 (22.5%) | 209 (100%) |  |
|  | Salaried employee | 19 (10.7%) | 92 (51.7%) | 67 (37.6%) | 178 (100%) |  |
| District of enrolment | Ranchi | 69 (12.2%) | 337 (59.5%) | 160 (28.3%) | 566 (100%) | **<0.001** |
|  | Godda | 59 (21.9%) | 182 (67.4%) | 29 (10.7%) | 270 (100%) |  |
|  | Saraikela | 45 (31.9%) | 81 (57.4%) | 15 (10.6%) | 141 (100%) |  |
| Monthly family income (in ₹) | <10000 | 99 (20.5%) | 318 (65.7%) | 67 (13.8%) | 484 (100%) | **<0.001** |
|  | ≥10000 | 74 (15.0%) | 282 (57.2%) | 137 (27.8%) | 493 (100%) |  |
| **Obstetric profile of study participants** |  |  |  |  |  |  |
|  | <3 | 53 (14.4%) | 204 (55.3%) | 112 (30.4%) | 369 (100%) | **<0.001** |
|  | ≥3 | 120 (19.7%) | 396 (65.1%) | 92 (15.1%) | 608 (100%) |  |
| Presenting trimester | First trimester | 22 (30.1%) | 45 (61.6%) | 6 (8.2%) | 73 (100%) | **<0.001** |
|  | Second trimester | 93 (22.8%) | 248 (60.8%) | 67 (16.4%) | 408 (100%) |  |
|  | Third trimester | 58 (11.7%) | 307 (61.9%) | 131 (26.4%) | 496 (100%) |  |
| Gravida | Primigravida | 97 (19.7%) | 309 (62.8%) | 86 (17.5%) | 492 (100%) | **0.018** |
|  | Multigravida | 76 (15.7%) | 291 (60.0%) | 118 (24.3%) | 485 (100%) |  |
| History of abortions | No | 146 (18.6%) | 498 (63.3%) | 143 (18.2%) | 787 (100%) | **<0.001** |
|  | Yes | 27 (14.2%) | 102 (53.7%) | 61 (37.1%) | 190 (100%) |  |
| **Laboratory and clinical findings** |  |  |  |  |  |  |
| Presence of anaemia based on Hb levels | No | 51 (16.9%) | 169 (56.1%) | 81 (26.9%) | 301 (100%) | **0.008** |
|  | Yes | 122 (18.0%) | 431 (63.8%) | 123 (18.2%) | 676 (100%) |  |
| Vitamin A deficiency | No | 170 (17.6%) | 592 (61.3%) | 203 (21.0%) | 965 (100%) | 0.513 |
|  | Yes | 3 (25.0%) | 8 (66.7%) | 1 (8.3%) | 12 (100%) |  |
| Iodine deficiency | No | 171 (17.6%) | 597 (61.4%) | 204 (21.0%) | 972 (100%) | 0.292 |
|  | Yes | 2 (40.0%) | 3 (60.0%) | 0 (0.0%) | 5 (100%) |  |
| Fluoride excess | No | 169 (17.5%) | 595 (61.8%) | 199 (20.7%) | 963 (100%) | 0.137 |
|  | Yes | 4 (28.6%) | 5 (35.7%) | 5 (35.7%) | 14 (100%) |  |
| Clinical anaemia | No | 126 (16.3%) | 463 (60.1%) | 182 (23.6%) | 771 (100%) | **<0.001** |
|  | Yes | 47 (22.8%) | 137 (66.5%) | 22 (10.7%) | 206 (100%) |  |

Among obstetric factors, number of ANC visits (p<0.001), trimester (p<0.001), gravida (p=0.018), and history of abortion (p<0.001) showed significant associations. (Table 2)

Anaemia based on Hb levels (p=0.008) and clinical anaemia (p<0.001) were also significantly associated with nutritional status. (Table 2)

### Multivariable analysis using multinomial logistic regression

After adjustment, age remained significantly associated with over nutrition. Women aged 25 to 29 years (aOR=5.26, p=0.001) and older than 29 years (aOR=6.44, p<0.001) had higher odds of over nutrition compared to those under 20 years. Participants belonging to Scheduled Tribe had significantly lower odds of overnutrition (aOR=0.33, p<0.001). Family incomes greater than ₹10,000 were associated with increased odds of over-nutrition. (aOR=1.87, p=0.007). Study participants recruited from Saraikela had higher odds of undernutrition (aOR=2.73, p<0.001). (Table 3)

**Table 3:** Multivariable analysis using multinomial logistic regression

| **Variables** |  | **Odds ratio (p-value)** |  | **Adjusted odds ratio (p-value)** |  |
|---|---|---|---|---|---|
|  |  | **Undernutrition vs Normal** | **Overnutrition vs Normal** | **Undernutrition vs Normal** | **Overnutrition vs Normal** |
| **Socio-demographic profile of study participants** |  |  |  |  |  |
| Age group | <20 years | 1 | 1 | 1 | 1 |
|  | 20-24 years | 0.82 (0.41) | **5.13 (<0.001)** | 1.13 (0.64) | **3.36 (0.015)** |
|  | 25-29 years | **0.49 (0.009)** | **9.61 (<0.001)** | 0.74 (0.35) | **5.26 (0.001)** |
|  | >29 years | 1.05 (0.88) | **11.15 (<0.001)** | 1.59 (0.22) | **6.44 (<0.001)** |
| Category | General | 1 | 1 | 1 | 1 |
|  | Scheduled Caste | 1.58 (0.17) | 0.37 (0.008) | 1.32 (0.43) | 0.46 (0.06) |
|  | Scheduled Tribe | 0.96 (0.87) | **0.27 (<0.001)** | 0.97 (0.93) | **0.33 (<0.001)** |
|  | Other Backward Classes | 0.68 (0.12) | **0.465 (<0.001)** | 0.65 (0.11) | **0.51 (0.004)** |
|  | Others | 1.12 (0.75) | 0.402 (0.010) | 1.18 (0.67) | 0.47 (0.06) |
| Educational status | Illiterate | 1 | 1 | 1 | 1 |
|  | Primary | 0.94 (0.82) | 1.34 (0.41) | 1.02 (0.95) | 1.10 (0.82) |
|  | Middle | 0.80 (0.44) | 1.57 (0.20) | 0.72 (0.33) | 1.12 (0.78) |
|  | Inter | 1.03 (0.93) | **2.47 (0.008)** | 1.17 (0.66) | 1.16 (0.71) |
|  | Graduate | **0.51 (0.045)** | **4.08 (<0.001)** | 0.67 (0.35) | 1.27 (0.58) |
| Occupation | Housewife | 1 | 1 | 1 | 1 |
|  | Household helper | 7.28 (0.11) | 2.99 (0.44) | 5.90 (0.18) | 8.53 (0.16) |
|  | Daily wage labour | **2.73 (0.026)** | 0.25 (0.18) | 2.51 (0.07) | 0.71 (0.75) |
|  | Business | 2.42 (0.17) | 0.50 (0.52) | 2.61 (0.18) | 0.54 (0.61) |
|  | Salaried employee | 0.36 (0.34) | **2.99 (0.016)** | 0.55 (0.60) | 1.19 (0.75) |
| Husband's Occupation | Unemployed | 1 | 1 | 1 | 1 |
|  | Daily wage labour | 2.31 (0.18) | 0.92 (0.85) | 2.08 (0.26) | 0.87 (0.80) |
|  | Business | 3.08 (0.08) | 1.61 (0.33) | 2.83 (0.12) | 1.25 (0.70) |
|  | Salaried employee | 1.65 (0.45) | **2.91 (0.027)** | 1.45 (0.61) | 1.30 (0.65) |
| District of enrolment | Ranchi | 1 | 1 | 1 | 1 |
|  | Godda | **1.58 (0.021)** | **0.336 (<0.001)** | 1.51 (0.21) | 0.55 (0.08) |
|  | Saraikela | **2.71 (<0.001)** | **0.390 (0.002)** | **2.73 (<0.001)** | **0.50 (0.035)** |
| Monthly family income (in ₹) | <10000 | 1 | 1 | 1 | 1 |
|  | ≥10000 | 0.84 (0.33) | **2.31 (<0.001)** | 0.89 (0.61) | **1.87 (0.007)** |
| **Obstetric profile of study participants** |  |  |  |  |  |
| Number of ANC checkups | <3 | 1 | 1 | 1 | 1 |
|  | ≥3 | 0.86 (0.41) | **2.363 (<0.001)** | 1.48 (0.095) | **2.11 (<0.001)** |
| Presenting trimester | First trimester | 1 | 1 | 1 | 1 |
|  | Second trimester | 0.77 (0.36) | 2.03 (0.12) | 0.67 (0.20) | 2.19 (0.10) |
|  | Third trimester | **0.386 (0.001)** | **3.200 (0.009)** | **0.27 (<0.001)** | 2.40 (0.076) |
| Gravida | Primigravida | 1 | 1 | 1 | 1 |
|  | Multigravida | 0.83 (0.29) | **1.457 (0.021)** | 0.87 (0.56) | 0.84 (0.45) |
| History of abortions | No | 1 | 1 | 1 | 1 |
|  | Yes | 0.90 (0.67) | **2.083 (<0.001)** | 1.16 (0.60) | **1.86 (0.009)** |
| **Laboratory and clinical findings** |  |  |  |  |  |
| Presence of anaemia based on Hb levels | No | 1 | 1 | 1 | 1 |
|  | Yes | 0.94 (0.74) | **0.595 (0.002)** | 0.77 (0.24) | 0.89 (0.56) |
| Clinical anaemia | No | 1 | 1 | 1 | 1 |
|  | Yes | 1.26 (0.24) | **0.409 (<0.001)** | 1.10 (0.73) | 0.70 (0.27) |

Among obstetric factors, third trimester was associated with lower odds of undernutrition (aOR=0.27, p<0.001). Having a history of abortion was significantly associated with higher odds of overnutrition (aOR=1.86, p=0.009). (Table 3)

No significant association was observed between anaemia and nutritional status after adjustment.

### Association of socio-demographic profile and dietary assessment with micronutrient deficiency or excess (fluoride)

Illiteracy, unemployment of husbands, and residence in Godda district were consistently associated with higher prevalence of anaemia, vitamin A deficiency, and iron deficiency, while daily wage labour was associated with vitamin A deficiency and fluoride excess. Tribal ethnicity and Christianity were significantly associated with anaemia. Younger age (<20 years) and primigravity were associated with iron deficiency, whereas age more than 29 years was associated with vitamin A deficiency. Fewer than three ANC visits were associated with anaemia.

Dietary factors also showed significant associations. Consumption of fewer than three meals daily was associated with anaemia, vitamin A deficiency, and iron deficiency. Lower intake of nuts and oilseeds was associated with anaemia, iron deficiency, and iodine deficiency, while lower intake of vegetables and fruits was associated with anaemia, vitamin A deficiency, fluoride excess, and iron deficiency. Lower intake of pulses and dairy products was associated with iron deficiency. Fluoride excess and iron deficiency were also more common among women consuming animal foods and vitamin A-rich foods at least twice weekly. Residence in Saraikela district was associated with higher iodine deficiency.

## Discussion

The present study demonstrates the coexistence of anthropometric malnutrition and micronutrient deficiency among pregnant women attending ANC clinics under PMSMA in Jharkhand, reflecting the double burden of malnutrition and hidden hunger during pregnancy. Based on BMI assessment, 38.6% of participants were malnourished, with 17.7% being underweight and 20.9% overweight or obese. Simultaneously, anaemia affected 69.3% of women, while clinical features suggestive of iron deficiency were observed among 21.1% of the participants. Vitamin A deficiency, iodine deficiency, and fluoride excess were also identified among a smaller proportion of women.

Dietary assessment revealed that cereals, millets, and pulses formed the staple diet, and regular intake of micronutrient-rich foods such as nuts and oilseeds, green leafy vegetables, fruits, and animal-source foods was inadequate among many participants. Such poor dietary diversity may partly explain the high burden of anaemia and micronutrient deficiencies observed in the study population.

The prevalence of anaemia in the present study was comparable to NFHS-5 estimates for Jharkhand. [19] Similar high burdens of anaemia among pregnant women have also been reported from Tamil Nadu, Maharashtra, and Nagaland. [11,22,23] The persistence of high anaemia prevalence despite ongoing supplementation programs indicates continuing gaps in maternal nutrition, dietary adequacy, and antenatal care services.

The prevalence of vitamin A and iodine deficiency in the present study was lower than that reported in previous studies. [23–26] This difference may be explained by the use of clinical assessment methods in the present study, which are less sensitive than biochemical investigations and may underestimate subclinical deficiencies.

This study also highlights the ongoing nutritional transition within Jharkhand. Rural districts such as Godda and Saraikela demonstrated higher burdens of undernutrition, whereas the more urbanized district of Ranchi showed higher levels of overnutrition. Similar rural-urban contrasts have been reported in NFHS-5 data and other studies. [19,27,28] These findings likely reflect differences in dietary patterns, socio-economic conditions, urbanization, and exposure to obesogenic environments.

The prevalence of undernutrition observed in the present study was comparable to findings reported from rural Karnataka, while studies from West Bengal demonstrated lower overall malnutrition burden. ^[29,30]^ The co-existence of undernutrition and overnutrition observed in the present study further supports the growing evidence of nutritional transition occurring in low- and middle-income countries. [31]

The strengths of the study include its large sample size, multi-centric design, and assessment of both anthropometric and micronutrient-related nutritional problems. However, the study also has certain limitations. Since the participants were selected by consecutive sampling, the study has a high risk of selection bias, negatively affecting the generalizability of the study. Several micronutrient deficiencies were assessed clinically rather than biochemically, which may have underestimated subclinical deficiencies. Dietary assessment was based on recall methods and may therefore be affected by recall bias. Additionally, the cross-sectional design limits causal inference.

## Conclusion

The findings of the present study have important public health implications. Despite the implementation of programs such as PMSMA, POSHAN Abhiyan and Anaemia Mukt Bharat, a considerable burden of maternal malnutrition and micronutrient deficiency persists. Strengthening routine nutritional screening, dietary counselling, and micronutrient supplementation during antenatal care, particularly among socio-economically vulnerable populations and women with poor ANC utilisation, remains essential.

Overall, the present study highlights substantial burden of both malnutrition and hidden hunger among pregnant women in Jharkhand and underscores the need for integrated maternal nutrition strategies during antenatal care.

## Data Availability

All data produced in the present study are available upon reasonable request to the authors.

